# Identifying alterations in hand movement coordination from chronic stroke survivors using a wearable high-density EMG sleeve

**DOI:** 10.1101/2024.01.02.24300714

**Authors:** Nicholas Tacca, Ian Baumgart, Bryan Schlink, Ashwini Kamath, Collin Dunlap, Michael Darrow, Samuel Colachis, Philip Putnam, Joshua Branch, Lauren Wengerd, David A Friedenberg, Eric C Meyers

## Abstract

Non-invasive, high-density electromyography (HD-EMG) has emerged as a useful tool to collect a range of neurophysiological motor information. Recent studies have demonstrated changes in EMG features that occur after stroke, which correlate with functional ability, highlighting their potential use as biomarkers. However, previous studies have largely explored these EMG features in isolation with individual electrodes to assess gross movements, limiting their potential clinical utility. Here, able-bodied (N=7) and chronic stroke subjects (N=7) performed 12 functional hand and wrist movements while HD-EMG was recorded using a wearable sleeve. We demonstrate that a variety of HD-EMG features, or views, can be decomposed from the wearable sleeve. Stroke subjects, on average, had higher co-contraction and reduced muscle coupling when attempting to open their hand and actuate their thumb. In an expanded dataset consisting of 37 movements, we characterized muscle synergies in the forearm of able-bodied individuals. We found that the high-density array provides additional resolution over manually placed electrodes, which may help dissociate finer nuances in motor control. Additionally, muscle synergies decomposed in the stroke population were relatively preserved, with a large spatial overlap in composition of matched synergies. Alterations in synergy composition demonstrated reduced coupling between digit extensors and muscles that actuate the thumb, as well as an increase in flexor activity in the stroke group. Average synergy activations during movements revealed differences in coordination, highlighting overactivation of antagonist muscles and compensatory strategies. When combining co-contraction and muscle synergy features, the first principal component was correlated with upper-extremity Fugl Meyer hand sub-score of stroke participants (R^2^=0.86). Principal component embeddings of individual features revealed interpretable measures of motor coordination and muscle coupling alterations. These results demonstrate the feasibility of predicting motor function through features decomposed from a wearable HD-EMG sleeve, which could be leveraged to improve stroke research and clinical care.

## Introduction

Successful execution of voluntary movement relies on a complex, dynamic relationship between the motor cortex, spinal cord, and skeletal muscles. This relationship is altered following a stroke, leading to impairment of various neural pathways responsible for healthy motor function. As a result, individuals recovering from stroke struggle with activities of daily living, including manipulation of objects such as doors, utensils, and clothing due to decreased upper extremity muscle coordination and weakness [1]. The heterogeneity in functional motor deficits among stroke survivors poses significant challenges for recovery and treatment, including the ability to assess treatment efficacy [2]. This variety in responses among individuals post-stroke creates the need for more sensitive measures of motor function.

Surface electromyography (EMG) provides a simple, non-invasive method to directly measure changes in muscle activation post-stroke. Simultaneous EMG recording from multiple muscles can reveal altered recruitment and coordination strategies across various movements. For example, co-contraction of the upper limb, which is characterized by overactivity in the antagonist muscles, is a common motor symptom following a stroke that is quantifiable using EMG sensors placed on the flexors and extensors of the forearm [3]. Some level of co-contraction during attempted movements can help stabilize the joint. However, excessive co-contraction of antagonist muscles can impair motor coordination, increase duration of movement, limit range of motion [4], lead to early onset fatigue [29], [30], and has been linked to the level of spasticity in stroke survivors [30]. Consequently, greater muscle spasticity is associated with greater motor impairment [5].

Similarly, muscle synergies, time-invariant patterns of muscle activations that produce controlled movement, and time-varying muscle synergy activations [6] provide a unique view into changes of muscle coordination after stroke. Compared to the non-paretic arm, the paretic arm often has a different number of active muscle synergies for a particular movement and a modified composition of muscle activation comprising those synergies [7]–[11]. Additionally, the ‘merging’ of synergies was observed after stroke, with the extent of merging reflective of the degree of residual motor functionality [12].

To date, collection of these and other EMG features have mainly been performed during gross movements using low-density bipolar EMG electrodes on a limited number of muscles. While this technology can capture data across several muscles, the electrodes are relatively large, preventing measurement of smaller hand muscles critical to properly perform functional tasks. In contrast, high-density EMG (HD-EMG) technology, which utilizes an array of densely packed electrodes, can provide a high-resolution view of motor control. The various features that can be extracted from HD-EMG may each capture unique aspects of motor control, and combinations could reveal new motor-related biomarkers after stroke. However, current HD-EMG systems are cumbersome and difficult to use, limiting their use to expert researchers.

In this work, we characterize movement coordination from the extrinsic hand muscles across a wide range of functional movements in both able-bodied and chronic stroke groups using an easy-to-use HD-EMG wearable sleeve that spans the forearm. First, we evaluate isolated control and muscle coupling via co-contraction index (CCI) and muscle correlation metrics, respectively. Next, we evaluate sub-muscle coupling to provide more detailed characterization of movement coordination via muscle synergy analysis across the forearm HD-EMG array. We compare muscle synergies decomposed from both groups to assess alterations in synergy composition that reflect functional differences. Lastly, we demonstrate the feasibility of combining HD-EMG features that correlate with functional movement ability in individuals with stroke. This work provides an initial demonstration of the wealth of information that can be extracted from a HD-EMG wearable sleeve, and preliminary identification of motor-related biomarkers that could be leveraged to improve stroke rehabilitation, movement disorders research, and clinical care.

## Materials and Methods

### Study Participants

Seven individuals (3 female, 4 male; 60±5 years) with a history of stroke and hemiparesis participated in the study. EMG was recorded from their affected arm using the NeuroLife® EMG sleeve [13] as they attempted various hand movements. An additional dataset of various movements and grasps was recorded from seven able-bodied (4 female, 3 male; 27±1 years) individuals to serve as a benchmark for comparison of HD-EMG features.

Both datasets were collected as part of an ongoing clinical study at Battelle Memorial Institute that was approved by the Battelle Memorial Institute Institutional Review Board (IRB0779 and IRB0773). All participants were informed of the study protocol and provided written consent prior to data collection in accordance with the Declaration of Helsinki. Participants with chronic stroke were eligible if they could follow three step commands, respond and communicate verbally, and their hemiparesis affected their arm and hand. Full inclusion/exclusion criteria are provided in the Supplementary Information.

Prior to EMG data collection, standardized clinical assessments were evaluated in participants with stroke and scored by a licensed occupational therapist. Assessments included the upper extremity section of the Fugl-Meyer (UEFM) evaluating gross arm movements and hand coordination sub-score (UEFM-HS), the Box and Blocks test to assess fine grip dexterity, and the Modified Ashworth Scale (MAS) test to assess finger and wrist spasticity. For some sub-analyses, participants were split into mild (UEFM-HS ≥ 3) and severe (UEFM-HS < 3) groups based on clinical assessments for group comparisons. Full participant demographics and clinical assessment data are contained in Supplementary Tables 1 and 2.

### Experimental Setup and Paradigm

The methods used in this experiment are similar to those previously described by Meyers et al. [13]. Participants were seated comfortably at a table with a computer monitor in front of them and both arms resting (Figure 1). Prior to donning the sleeve, the forearm was sprayed with an electrode solution spray (Signaspray, Parker Laboratories, Fairfield, NJ) to enhance signal quality. The NeuroLife EMG sleeve was then donned on the affected arm for individuals with stroke and on the right arm for able-bodied individuals. The sleeve consists of a stretchable lightweight fabric with embedded electrodes that record bipolar EMG. The sleeve comes in three different sizes: small: 128 electrodes (59 channels), medium: 142 electrodes (70 channels), and large: 150 electrodes (75 channels). Each subject’s sleeve size was determined based on forearm size and comfortability once donned (Supplementary Tables 1 and 2).

**Figure 1.**
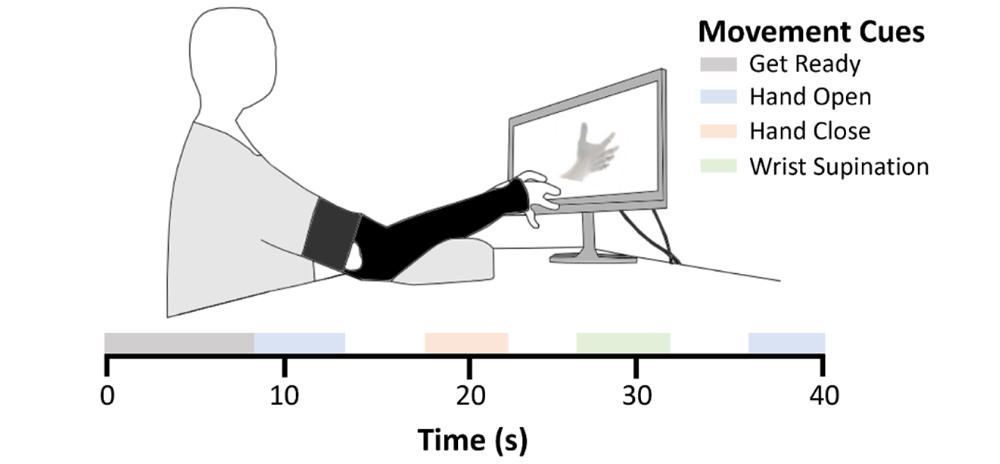
Illustration of experimental data collection procedure. Participants were seated across from a computer monitor that displayed sequential cues (e.g., Hand Open, Hand Close, Wrist Supination) to follow. The HD-EMG sleeve was donned on th affected arm for participants with stroke and on the right arm of able-bodied participants. Participants attempted the cue movements to the best of their ability at approximately 25-50% MVC effort. An operator monitored the data collection to ensure adequate signal and participant engagement.

Pictorial cues of various hand, wrist, and forearm movements were displayed on the monitor for participants to follow. Participants were instructed to attempt movements to the best of their ability at approximately 25-50% of their maximal effort level regardless of their ability to physically perform the cued movement. Based on participant preference, an optional foam cushion was used to prop up the arm used to attempt the movements.

Both stroke and able-bodied participants attempted the following 12 movements: Hand Close, Hand Open, Pointing Index, Thumb Flexion, Thumb Extension, Thumb Abduction, Wrist Supination, Wrist Pronation, Wrist Flexion, Wrist Extension, Thumb Two Point Pinch, and Key Pinch. These movements have been used in previous studies [13], [14] due to their relevance for dexterous hand use in functional tasks. An additional 25 different movements and fine grasps (for a combined total of 37 movements) were performed by able-bodied participants to fully characterize coordination across a wide range of movements and grasps (Supplementary Figure 9). These 25 movements were not collected in stroke participants because they consisted of highly dexterous movements that stroke participants could not perform.

We recorded HD-EMG in blocks that consisted of individual movements repeated, as well as mixed blocks with up to three different movements repeated. Both block types began with an 8-second “Get Ready” period prior to showing any cues, and a rest period was always interleaved between all cued movements. Each cue was prompted for 4-6 seconds for individuals with stroke to account for slower reaction times, and 2-3 seconds for able-bodied participants. Cue duration was selected randomly from a uniform distribution within each group’s respective cue duration ranges. The stroke movement dataset was collected in a single session lasting no more than two hours. Each participant attempted each movement at least 15 times. For the able-bodied dataset, EMG was recorded in a single session in which each movement and grasp was performed at least 10 times.

### EMG Signal Processing

EMG data were recorded at a sampling rate of 3,000Hz using an Intan Recording Controller (Intan Technologies, Los Angeles, CA). Pre-processing consisted of notch filtering the data at 60 Hz and bandpass filtering between 20 and 400 Hz with a 10^th^ order Butterworth filter [13], [15]. Prior to EMG feature extraction, additional artifact filtering steps were performed to correct artifacts due to sleeve shift, electrode lifting, and impedance changes over time. A blind source separation (BSS) technique based on the approximate joint diagonalization of cospectral (AJDC) covariance matrices adapted from the open-source pyRiemann implementation [16] based on [17], [18] was used to find spectrally uncorrelated sources. The open-source QNDIAG algorithm [19] was used to jointly diagonalize the cospectral covariance matrices, as it exploits a state-of-the-art quasi-Newton strategy for an increase in computation speed and separability between sources in the spectral domain. Using kurtosis with a threshold of 95%, artifact sources were automatically detected in the source signal and suppressed from the reconstructed signal. Figure 2A shows the final filtered EMG signal in three representative channels. Supplementary Figure 2 gives a stepwise filtering example using the BSS AJDC method for artifact detection and correction. Following artifact correction, the root mean square (RMS) of 150ms bins with stride of 40ms was used for subsequent feature decomposition steps (Figure 2B). The full EMG signal processing pipeline from raw data to feature decomposition from a representative stroke subject is shown in Figure 2. Refer to Supplementary Figure 1 for a comparable pipeline from an able-bodied subject.

**Figure 2.**
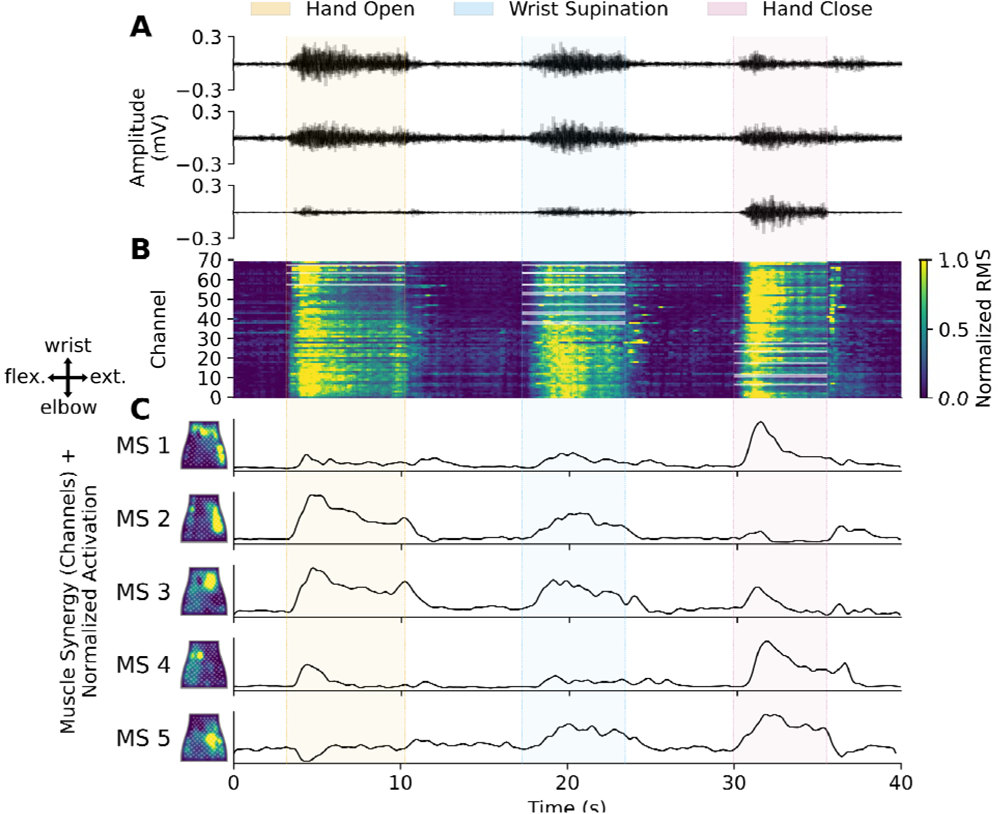
Representative pipeline from raw HD-EMG signal to feature decomposition across three cued movements from an individual with chronic stroke. **(A)** Filtered EMG from three representative channels recorded using the NeuroLife sleeve. **(B)** Time-series heatmap of the globally normalized root mean square (RMS) of 150ms bins with stride of 40ms of EMG activity. Highlighted channels during the cued movements correspond to the agonist channels defined in the muscle group masks (Supplementary Figure 4 and Supplementary Table 3). (C) *Left:* Heatmaps of time-invariant muscle synergies decomposed with non-negative matrix factorization (NMF) mapped to the flattened HD-EMG sleeve. Active areas correspond to the weighting of coupled muscles for each muscle synergy. A compass showing the orientation of the sleeve mapping is shown on the left (flex. = flexors, ext. = extensors). *Right:* Activation of the muscle synergies across time during the different attempted movements.

### EMG Sleeve Mapping

#### Mapping between sleeve sizes and arms

To compare decomposed HD-EMG features across subjects with different sleeve sizes and arms, a standardized sleeve mapping procedure was conducted (Supplementary Figure 3). Once the windowed EMG data were processed into RMS features, all sleeve sizes were mapped using linear interpolation to the medium sleeve for a total of 70 feature channels. HD-EMG data were only recorded from the left arm of one subject (Subject 3, stroke group). In this instance, the channel data were mirrored to map to the right arm. The medium sleeve has a direct mirroring except for the ground electrode in the bottom left corner. An additional linear interpolation was used to account for the ground electrode mirroring. The final output of the sleeve mapping procedure ensured all EMG feature data were mapped to the medium sleeve as if it was worn on the right arm for consistent comparisons between subjects. This procedure was conducted prior to co-contraction and muscle correlation analyses for mapping to muscle groups described in the following section. For muscle synergy analysis, muscle synergies were first decomposed in the original sleeve arm and size to retain the full signal resolution during decomposition, with the sleeve mapping procedure occurring post-decomposition.

#### HD-EMG to muscle group mapping

To map the HD-EMG sleeve to muscle groups for selecting agonist muscles during movements for co-contraction analysis and physiological interpretation of EMG feature results, a HD-EMG-to-muscle group mapping was developed for the medium sleeve worn on the right arm based on the able-bodied dataset. We selected movements actuated by a single agonist muscle or muscle group (e.g., Hand Close is accomplished through actuation of the digit flexor muscle group; Supplementary Table 3). The agonist muscle group, therefore, was the predominant source of EMG activity during each selected movement. HD-EMG recordings for these movements were averaged within session and across all participants. These averages were converted to binary masks across EMG channels with a 95% amplitude threshold for the following muscle groups: Digit flexors, Digit extensors, Wrist flexors, Wrist extensors, Thumb extensors, and Thumb flexors (Supplementary Figure 4). This HD-EMG channel-to-muscle mapping scheme was validated through a leave one out (LOO) cross validation showing that 86.9 ± 0.84% of single agonist movement activity was captured by the masked data. When mapping to muscle groups, the full channel-wise data was multiplied with the muscle masks and averaged based on the number of channels that contribute to each mask. This reduced the channel data down to the six muscle groups representative of manually placed EMG electrodes on the forearm.

### HD-EMG Feature Decomposition

#### Co-contraction index and muscle coupling

A simple co-contraction index (CCI) based on ref [20] was used to benchmark the amount of co-contraction of non-agonist muscles during individual movements:

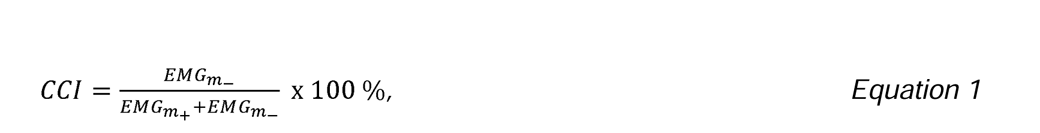

where *CCI* is the co-contraction index, *EMG_m__*represents EMG data from the non-agonist muscle groups, and *EMG_m+_*represents EMG data from the agonist or targeted muscle group during movement. Corresponding agonist and non-agonist muscle groups were determined via a HD-EMG-to-muscle-group mapping based on average able-bodied muscle masks during cued movements (Supplementary Figure 4 and Supplementary Table 3). Muscle coupling was assessed using average Pearson correlation between muscle groups. Correlations between muscle groups were z-transformed prior to averaging and then reconstructed post-averaging for group results.

#### Muscle synergies

For a more comprehensive assessment of movement coordination, non-negative matrix factorization (NMF) [21] from the Scikit learn toolbox [22] was used to decompose the globally normalized RMS feature data into time-invariant muscle synergies and time-varying synergy activations (Figure 2C). This matrix factorization method was chosen due to its robustness across datasets [23]. The number of muscle synergies used to describe the data sufficiently was based on an 85% variance accounted for (VAF) threshold between the reconstructed data and the original signal [7], [24]. Muscle synergy components were ordered according to the average across participants’ individual synergy component reconstruction (VAFi). Next, muscle synergies by channels were mapped to the medium sleeve as if it were worn on the right arm for comparisons between subjects and for the HD-EMG channel-to-muscle mapping. Once mapped to the medium-right sleeve setting, muscle synergies were reduced to muscle group weightings via the HD-EMG to muscle groups mapping for additional physiological interpretation. Lastly, average muscle synergy activation over individual movements was determined to map the synergy weighting to the different movements.

### HD-EMG Feature Analysis

To compare HD-EMG features between subjects with different sleeve sizes and arm recordings, all recordings and muscle synergies were mapped to the medium-right sleeve configuration using the sleeve mapping procedure previously described. Average CCI and muscle correlation by movement were compared between groups with paired t-tests. For muscle synergy analysis, a LOO analysis was conducted in which a single subject’s extracted muscle synergy components were compared to the remaining group average. Cosine similarity was used as the metric for determining muscle synergy similarity between subjects and group averages [25]. When decomposing to more than one muscle synergy, the median cosine similarity of each mapped muscle synergy comparison was reported. Muscle synergies were mapped across individuals by ordering the synergies based on cosine similarity.

A group principal component analysis (PCA) was conducted on a HD-EMG feature set containing CCI by movements and muscle synergies by channels across subjects [26], [27]. First, PCA was performed on the concatenated feature set to predict clinical UEFM-HS scores from the combined first principal component using linear least squares regression. Next, individual projections for each feature were computed with least squares estimation to map each feature to the individual reduced subspace principal components. Subsequent individual feature embeddings to transform the reduced subspace back to the original feature set were obtained by taking the pseudo-inverse of the individual projections. The feature embeddings highlight weightings to each individual feature to help with interpretability of the reduced principal components. Lastly, principal components and embeddings were inverted for plotting visualization.

### Statistical Analysis

Comparisons conducted were a priori. Lillifors tests were performed to test normality of distributions. Paired t-tests were used to determine significant differences between groups in both LOO analysis and between groups analysis. An alpha value of 0.05 was used for single comparisons. All statistical analyses were performed using Python 3.8 using SciPy [28]. For all figures, * indicates p<0.05, ** indicates p<0.01, and *** indicates p<0.001. Error bars in figures indicate mean ± standard error of the mean (SEM).

## Results

### Alterations in isolated control of the hand extrinsic muscles after stroke

Using our HD-EMG sleeve, we found that stroke subjects have more co-contraction relative to able-bodied subjects in movements that actuate the thumb, digit extensors, and wrist supinator muscles (Figure 3A and Supplementary Figure 5). Specifically, CCI was higher for Hand Open (p=0.029), Thumb Flexion (p=0.006), Thumb Extension (p=0.014), Thumb Abduction (p=0.017), and Wrist Supination (p=0.010) for individuals with stroke. We then assessed the relationship between muscle groups via a muscle coupling network. Able-bodied subjects had greater coupling between muscles than stroke subjects based on the grand average Pearson muscle correlations (Figure 3B), suggesting decreased motor recruitment during select movements after stroke. Full CCI and Pearson correlation results for the remaining movements are shown in Supplementary Figure 5.

**Figure 3.**
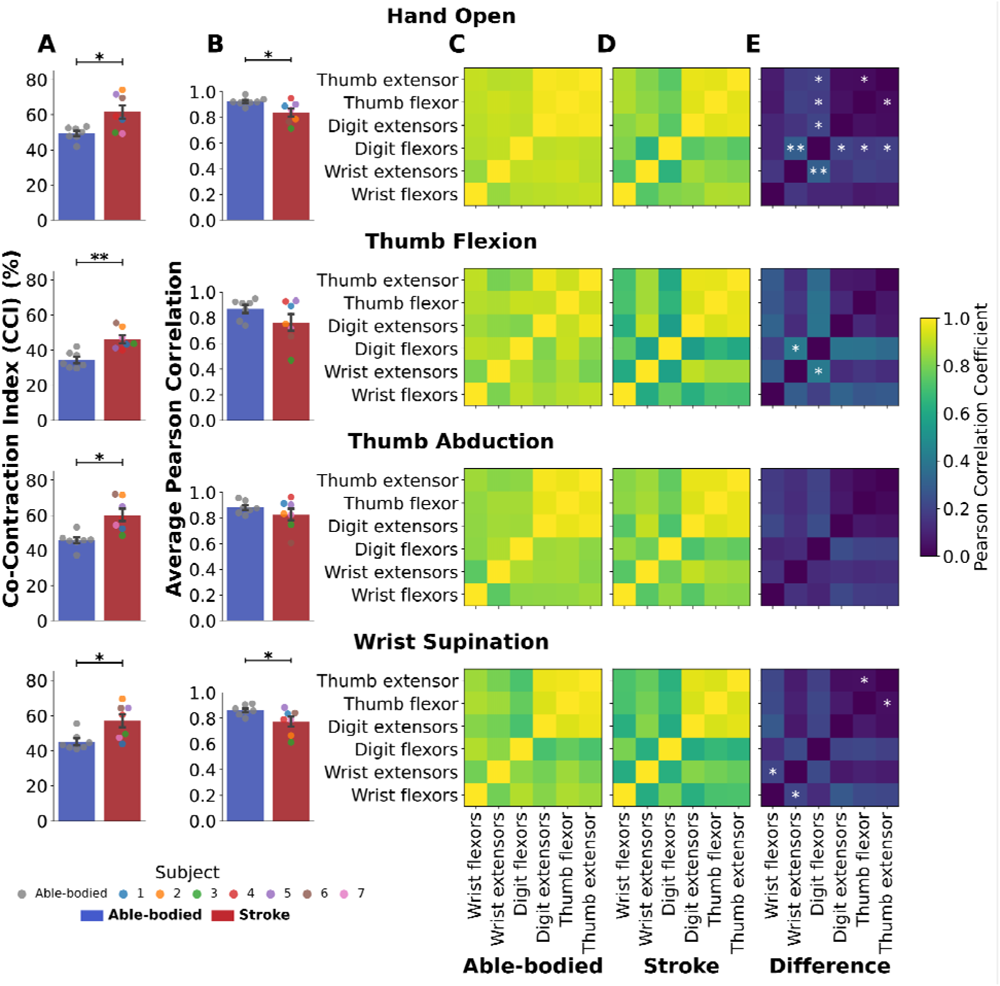
Co-contraction and muscle coupling across muscle groups during four attempted movements. **(A)** Co-contraction index (CCI) and **(B)** average Pearson correlation between muscle groups for able-bodied (blue bars) and stroke (red bars) groups. CCI was greater in the stroke group across all four movements. A higher muscle coupling was observed in able-bodied participants for Hand Open and Wrist Supination. **(C-E)** Average correlation matrices between muscle groups across able-bodied, stroke, and the difference between the two groups show able-bodied subjects have more coupling between certain muscle groups. Coupled muscle groups determined with paired t-tests are denoted with * (p<0.05) and ** (p<0.01) indicating significant differences betwee groups.

Muscle correlation networks revealed reduced coupling between digit flexors and wrist extensors (p=0.006), digit extensors (p=0.022), thumb flexor (p=0.046), and thumb extensor (p=0.041) muscle groups in the stroke group (Figure 3C-E). Similarly, thumb muscles were more coupled for able-bodied participants than participants with stroke for Hand Open (p=0.022) and Wrist Supination (p=0.047) movements. Digit flexors and wrist extensors, as well as wrist extensors and flexors were more coupled in able-bodied individuals for Thumb Flexion (p=0.025) and Wrist Supination (p=0.050), respectively. Together, these results highlight the ability to extract precise information related to hand extrinsic muscle coupling of individuals with stroke using a HD-EMG sleeve.

### HD-EMG sleeve provides high-resolution muscle synergies

While creating networks of correlated muscles can assess coupling during specific movements, muscle synergy analysis can provide a more holistic view of synergistic muscles across a wide range of movements. To benchmark coordination of hand movements in the able-bodied group, we decomposed the HD-EMG signals to time-invariant muscle synergies by channels (Figure 4A) and by muscle groups (Figure 4B), as well as time-averaged synergy activations across movements (Figure 4C). Across the seven able-bodied subjects, seven muscle synergies were able to reconstruct the data with at least 85% explained variance (Figure 4D). Synergies were ordered based on the average individual explained variance when reconstructing the original signal (Figure 4E).

**Figure 4.**
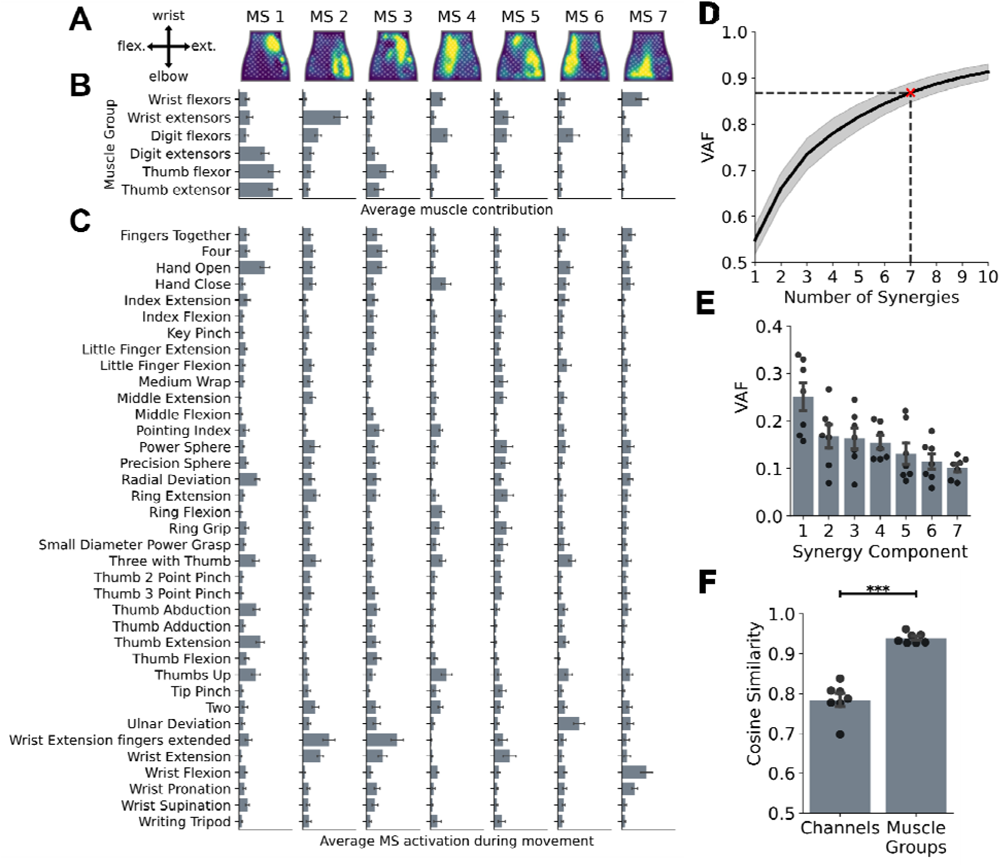
Muscle synergies decomposed in the forearm of able-bodied (N=7) subjects that performed 37 different movements and grasps. **(A)** Average time-invariant muscle synergy (MS) representations by EMG channels across all subjects mapped to the flattened NeuroLife sleeve. Active regions highlight coupled forearm muscles. **(B)** Average time-invariant muscle synergy representations mapped to muscle groups across all subjects. **(C)** Average activation of muscle synergies during the cued movements. **(D)** Average variance accounted for (VAF) between the original and reconstructed signals ± standard error of the mean. Seven muscle synergies were sufficient to reconstruct the data with at least 85% of the explained variance. **(E)** Individual VAF for each synergy component. Note: synergy components are ranked in order of explained variance of the data. **(F)** Leave one out (LOO) analysis comparing each subject’s muscle synergies with the remaining group average. Each point represents th median cosine similarity between each muscle synergy pair of the seven muscle synergies. The average cosine similarity across the seven muscle synergies is shown for both the channels (A) and muscle groups (B) muscle synergy representations. Using the reduced muscle synergy representation via muscle groups, similar to using single electrodes, the cosine similarity is significantly higher among individuals indicating that the HD-EMG array provides additional resolution to dissociate movement coordination (p < 0.001).

The first muscle synergy (Figure 4A – MS 1) was composed of digit extensors and muscles that actuate the thumb. This synergy was prominently activated during Hand Open, as well as various thumb movements and grips. Aside from the coupling with muscles that actuate the thumb in the first synergy, digit extensors were not a prominent component in any of the other synergies decomposed. The second muscle synergy consisted largely of the wrist extensors with some coupling to the digit flexors as well for stabilization during Wrist Extension movements. Interestingly, the third synergy consisted largely of muscles that actuate the thumb only. This synergy was also activated during Wrist Extension movements. Visually, the fourth and sixth synergies appear similar based on the heatmap representation, though the fourth synergy weighted muscles slightly more in the medial-distal region of the sleeve. However, while both were composed of digit flexor muscles and were activated during Hand Close, contrary to the fourth synergy, the sixth synergy was activated more during Ulnar Deviation. The fifth synergy was similar to the second synergy, albeit with a nearly equal weighting of wrist extensors and digit flexors. It also appears to have been activated during similar movements to the second synergy. Finally, the seventh synergy largely consisted of wrist flexors and was subsequently activated during Wrist Flexion and Wrist Pronation movements.

To compare synergies between subjects, a LOO analysis was conducted in which each subject’s synergies were compared with the remaining group average. Each synergy was mapped to the group average based on highest degree of similarity across all seven synergies. The median cosine similarity between matched synergies is shown as an individual subject data point in Figure 4F for both the channel (Figure 4A) and muscle group (Figure 4B) synergy representations. Muscle synergies mapped to muscle groups, representative of individually placed electrodes on forearm muscle groups, were more similar between subjects with an average cosine similarity of 0.938 ± 0.005, with the full channel representation at 0.784 ± 0.017 similarity across subjects (p < 0.001). This supports the notion that a high-density array provides additional resolution over manually placed electrodes, which may help dissociate finer nuances in movement coordination.

### Muscle synergies are relatively preserved following stroke, with alterations highlighting specific movement coordination differences

In the 12-movement dataset, muscle synergies and synergy activations were decomposed from both able-bodied and stroke populations to investigate potential changes in movement coordination post-stroke (Figure 5). With the downselected dataset, only five synergies were required to reconstruct the original signal in both groups (Figure 5D). Overall, muscle synergies decomposed from the channel data match closely between the stroke and able-bodied groups, though the spatial activation of the stroke group’s synergies are slightly more distributed across the sleeve (Figure 5A). Average group synergies by channels and synergy activations by movements over time can be seen in Supplementary Figures 6 and 7. When looking specifically at muscle groups, the coupling between stroke subjects’ digit extensors and muscles that actuate the thumb was affected in the first synergy (Figure 5B – MS 1). This synergy was still activated during Hand Open in both groups, but it was also largely activated during Hand Close, Key Pinch, Pointing Index, Wrist Pronation, and Wrist Supination for the stroke group and not in the able-bodied group (Figure 5C). Instead, the second synergy, which favored wrist extensors in both groups, had the highest activation for Hand Open in the stroke group, with the able-bodied group only having a moderate activation. This demonstrates a compensatory strategy in which stroke individuals favored wrist extensors over digit extensors when attempting to open their hands. Interestingly, synergy 5, which was composed of wrist flexors in both groups, had a more dominant weighting for stroke subjects compared to the able-bodied group. This synergy was also highly activated in both Wrist Flexion and Extension, as well as some other movements, in the stroke group only. This demonstrates an excessive co-contraction of the wrist flexor muscles during attempted movements that is not present in the able-bodied group. This can also be seen in the activation of synergy 4, which favored the digit flexors strongly in both groups.

**Figure 5.**
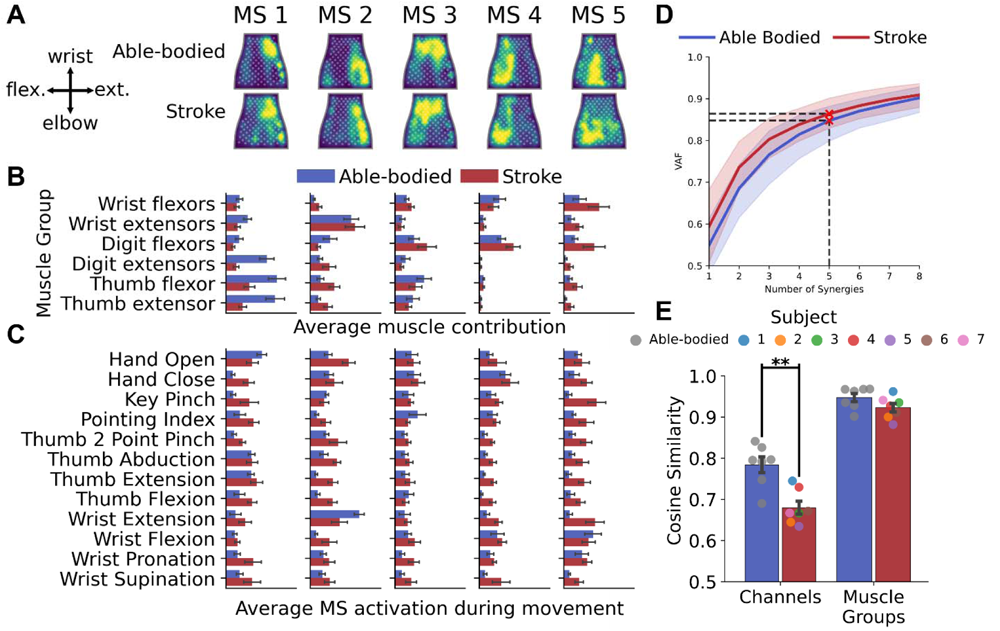
Muscle synergies decomposed in the forearm of able-bodied (N=7) and chronic stroke (N=7) subjects from various functional movements. Muscle synergies are ordered based on the individual variance accounted for (VAFi) when reconstructing the original signal. **(A)** Average time-invariant muscle synergy (MS) representations by EMG channels across all subjects within groups mapped to the flattened NeuroLife sleeve. On average, muscle synergies in the stroke group are similar to the average able-bodied muscle synergies, indicating that muscle synergies are relatively preserved following a stroke. **(B)** Average time-invariant muscle synergy representations mapped to muscle groups across all subjects within groups. On average, able-bodied subjects had a higher weighting to digit extensors and thumb muscles in the first muscle synergy. **(C)** Average synergy activations during the attempted movements. **(D)** Average variance accounted for (VAF) between the original and reconstructed signals ± standard error of the mean. Five muscle synergies were sufficient to reconstruct the data with at least 85% of the explained variance for both groups. **(E)** Leave one out (LOO) analysis of muscle synergy similarity. Each point represents the median cosine similarity between a subject’s muscle synergies and the remaining group average. On average, muscle synergies in their channel representation (A) were more similar within able-bodied subjects than within stroke subjects (p < 0.01), whereas there was no significant difference when mapped to muscle groups analogous to manually placed EMG electrodes.

To assess muscle synergy similarity between groups, a LOO analysis was conducted (Figure 5E). When decomposed in their full HD-EMG channel representation (Figure 5A), muscle synergies were more similar within able-bodied subjects than within stroke subjects (p < 0.01). This demonstrates the variability between participants with stroke, which may be associated with differing functional abilities. However, when reducing muscle synergies to muscle groups, analogous to individually placed EMG electrodes, there was no significant difference between groups. As the spatial resolution decreased, the synergies matched more closely between subjects and between subject groups, indicating there may be some additional sub-muscle coupling [29]. Supplementary Figure 8 shows individual subject comparisons to the average able-bodied control group’s synergies by channels.

### Interpretable, combined measures of movement coordination can predict functional clinical metrics

To demonstrate the potential for HD-EMG features to predict motor function, we used a combinatorial approach in which multiple feature views were assessed. Taking into consideration both co-contraction by movement and time-invariant muscle synergies can help assess motor control from two related perspectives. A concatenated HD-EMG feature vector was created consisting of CCI by movement (12 movements) and stacked muscle synergies by channels (5 synergies x 70 channels) for a total of 362 features per subject. This feature vector was reduced using PCA to its first two combined principal components explaining 47.0% (PC1) and 16.5% (PC2) of the data, respectively. Individual feature projections were determined using least squares estimation [26] to map the features to individual principal components (Figure 6A). Based on these two components, three groups of distinct functional ability emerged (able-bodied, UEFM-HS > 3 – mild, UEFM-HS ≤ 3 – severe). Additionally, the first principal component was able to predict UEFM-HS in stroke subjects with R^2^=0.86 (p=2.53 x 10^-3^) (Figure 6B).

**Figure 6.**
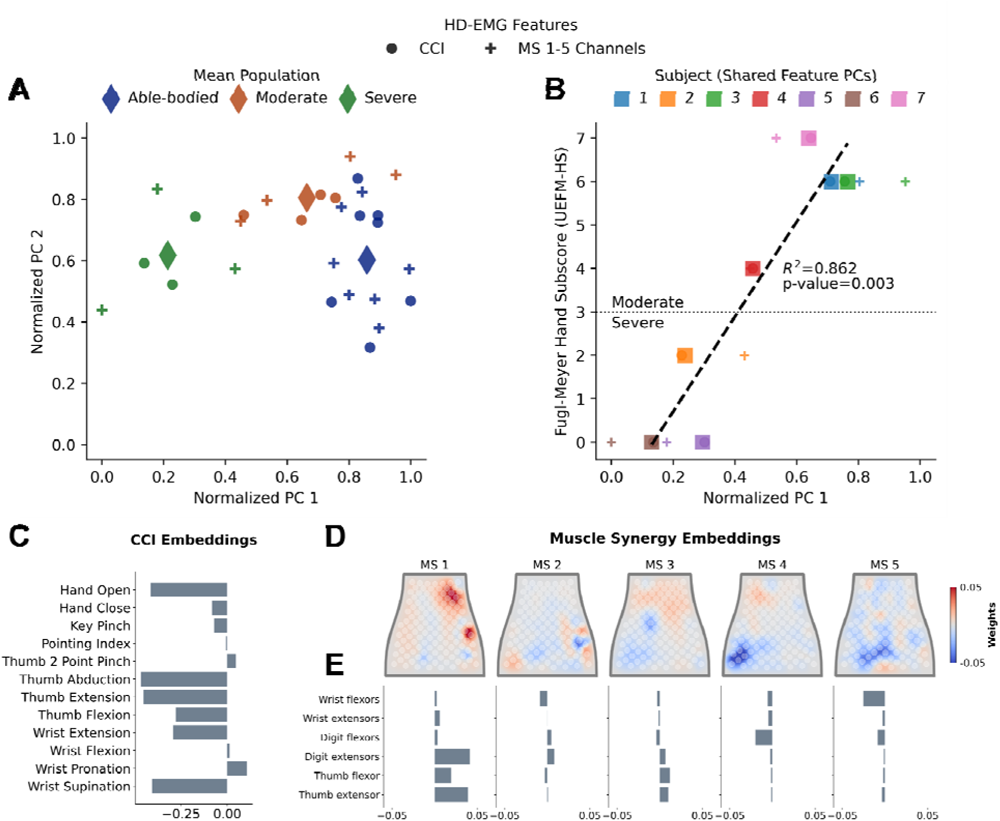
Combining average co-contraction by movement and muscle synergies by channels to predict groups of functional ability based on the Upper Extremity Fugl-Meyer – Hand Subscore (UEFM-HS) clinical assessment. **(A)** Scatter plot of the first two principal components of the HD-EMG features highlighting three distinct populations (Able-bodied, mild: UEFM-HS > 3, and severe: UEFM-HS ≤ 3). The diamonds indicate the population average. **(B)** The shared first principal component is correlated with UEFM-HS in stroke participants R^2^=0.86 (p=2.53 x 10-3). **(C)** Individual embeddings of the average co-contraction by movement show that a negative weighting, or a reduction in CCI, for Hand Open, Thumb movements, Wrist Extension, and Wrist Supination explains a positive increase in functional ability. **(D, E)** Individual embeddings of muscle synergies by channels mapped to the flattened sleeve (D) and muscle groups **(E)**. An increase in coupling between the digit extensor and muscles that actuate the thumb in the first synergy explains an increase in functional ability by clinical score. Alternatively, a negative weighting in digit flexors and wrist flexors in synergies 4 and 5, respectively, demonstrates increased flexor activation for more severe subjects.

Interpretability of alterations in biomarker features is important to understand deficiencies, which may reveal mechanisms of recovery and potentially new therapies. By looking at the first principal component embeddings of individual features, it is possible to see affected movements and alterations in muscle coupling. Individual principal component embeddings mapped to CCI by movement show a strong negative weighting for Hand Open, Thumb movements, Wrist Extension, and Wrist Supination indicating that as CCI decreases, functional ability improves (Figure 6C). Embeddings mapped to muscle synergies show a positive weighting to the digit extensors and muscles that actuate the thumb in the first muscle synergy (Figure 6D & E). This demonstrates that an increase in functional ability based on clinical score can be explained by an increase in coupling between digit extensors and muscles that actuate the thumb. On the other hand, a negative weighting in digit flexors and wrist flexors in synergies 4 and 5, respectively, demonstrates increased flexor activation for more severe subjects.

## Discussion

In this study, we demonstrated the ability to characterize movement coordination across a wide range of hand movements using a HD-EMG sleeve that spans the forearm muscles. Our results build upon previous studies that assessed gross arm movements by showing that synergies of the forearm during dexterous movements are also relatively preserved in composition following cortical stroke [7], with changes in composition related to affected muscle coupling that hinders functional ability [30]. We show that activation of synergies during movements revealed differences in coordination, highlighting overactivation of antagonist muscles and compensatory strategies. By developing a mapping between sleeve sizes, we were able to compare subjects of varying functional ability and arm sizes with a streamlined sleeve system that can be easily adopted by both research and clinical communities.

To provide physiological interpretation of the extracted HD-EMG features, we developed a HD-EMG-to-muscle group mapping. This mapping highlighted reduced coupling between digit extensors and muscles that actuate the thumb in the chronic stroke population in both muscle correlation and muscle synergy analyses. This reduction in muscle coupling was associated with a reduction in functional movement ability (Figure 6B). Despite the reduced coupling between these muscle groups, stroke subjects had greater overall co-contraction than able-bodied subjects during select movements (Figure 3A). Muscle synergies represented in their reduced spatial mapping to muscle groups showed a higher similarity between subjects than the channel-wise muscle synergy representation (Figure 4F). This suggests that manually placed electrodes on a small number of prominent muscles may not fully characterize differences between synergies across a wide range of dexterous movements. Improved spatial coverage provided by the sleeve’s HD-EMG array spans multiple muscles and can extract common synaptic input to muscles that contribute to multiple synergies [31], [32]. In a similar study, researchers used HD-EMG to account for subtle variation of neuromuscular activities via muscle synergies across channels during two wrist movements under isometric and isotonic training modes [29]. Our results further support the finding that whole-muscle components of synergies can be broadened to include sub-muscle components represented by HD-EMG. Furthermore, our work expands this finding to account for a full range of functional hand movements, as well as fine grasps, in both an able-bodied and stroke population to characterize hand movement coordination. By evaluating both the full and reduced muscle synergy representations, subtle changes in muscle activation can be assessed and muscle groups with reduced coupling can be targeted for further investigation.

Singular features of HD-EMG may not be able to fully characterize an individual’s level of motor function. As a result, we assessed a combination of HD-EMG features (CCI and muscle synergies) to predict functional ability measured with standard clinical assessment metrics. We showed that a single principal component from the combined feature vector is correlated with UEFM-HS in stroke subjects with varying levels of motor ability. These results demonstrate that features decomposed from HD-EMG can dissociate varying abilities to perform functional hand movements.

By characterizing alterations in HD-EMG features post-stroke, we may be able to identify candidate biomarkers of motor function across several categories that can be leveraged to ultimately improve functional outcomes. Evaluation of new therapies remains difficult due to unpredictable, “spontaneous” recovery experienced by stroke, thus creating a challenge for clinical trials evaluating restorative and rehabilitation interventions because it introduces significant variance [33]. To this end, a prognostic biomarker to predict recovery potential could be used to stratify participants to reduce inter-group variability, thereby increasing statistical power and reducing sample size, cost, and study durations. Assessment of upper limb function and recovery progress is typically measured using validated clinical measurement tools, including the Action Research Arm Test (ARAT) and UEFM scale. While these tests provide sensitive measures of function [34], [35], they can take anywhere from 10-20 minutes to administer, decreasing their clinical feasibility. A monitoring biomarker comprised of real-time HD-EMG features, such as changes in muscle synergies across task practice during rehabilitation, could lead to more informed treatment plans that target specific motor deficiencies. Finally, several neurological disorders present with overlapping motor symptoms that pose challenges for early diagnosis [36]. Comprehensive assessment of features derived from HD-EMG may be able to produce a diagnostic biomarker that can differentiate between these disorders.

We demonstrated that HD-EMG features were correlated with motor function in chronic stroke survivors, but the methods described have the potential to be broadly applied across a wide range of applications. Our analysis focused on movement coordination of the hand, but lower limb EMG patterns post-stroke further demonstrate the heterogeneity across individuals [37], presenting an opportunity to leverage the spatiotemporal advantages of HD-EMG to classify motor function across patients. HD-EMG features may also be able to provide insights into altered motor function of other neurological disorders, including Parkinson’s disease, dystonia, and amyotrophic lateral sclerosis (ALS). Our preliminary analyses suggest that combining HD-EMG features into an interpretable metric is useful for predicting motor function. It is likely that different sets of HD-EMG features and clinical information may provide unique prediction capabilities for each disorder.

While we successfully demonstrated the ability to characterize hand movement coordination in both able-bodied and stroke populations, we extracted HD-EMG features across simple movements. Future studies should consider compound movements to better approximate activities of daily living (e.g., grasping with supination in a bottle pour task). Despite focusing on simple movements, co-contraction and muscle synergies were able to dissociate functional ability between varying levels of stroke survivors. It is possible that over the course of therapy, however, minor changes in HD-EMG features may not directly correlate with functional improvement. To uncover subtle changes, in future studies, we should further leverage the HD-EMG array signal resolution to assess the neural drive to muscles via motor unit decomposition to assess the common input to motor neurons for modular control [31], [32], [38]–[40]. Using a compound movement dataset and combining different aspects of motor control through a single system can help assess motor function with a more holistic framework. Additionally, while we were successfully able to compare features extracted between subjects with different arm sizes and anatomy, it would be beneficial to record EMG from the unaffected arm of hemi-paretic stroke participants for a more direct comparison of recovery. This could also refine a synergy-based functional electrical stimulation (FES) pattern focused on alterations in synergy composition, which has shown promise to promote recovery in stroke individuals’ upper-limb motor performance [41]–[45]. Incorporating FES within the same HD-EMG recording electrodes in a single sleeve formfactor may help to personalize FES based on EMG features. Our group is actively developing an EMG-FES sleeve system to provide targeted stimulation based on motor intention to drive neuroplasticity.

## Conclusion

In this study, we successfully demonstrated that HD-EMG features decomposed from the paretic arm of chronic stroke survivors show changes in movement coordination that correlate with motor function. We demonstrated the ability to compare sub-muscle synergies across individuals and groups to highlight alterations that reflect differences in muscle coupling during hand movements. Muscle synergies that coordinate hand movements were relatively preserved following cortical stroke, with reduced coupling observed in the digit extensors and muscles that actuate the thumb, as well as an overactivation of flexors. Importantly, we introduced the concept of combining different HD-EMG features of motor control for a more holistic view of movement coordination and functional ability. We found that there was an interpretable combination of HD-EMG features that was correlated with clinical scores of motor function. Overall, these findings suggest that non-invasive HD-EMG features can describe altered motor control post-stroke, providing an opportunity to leverage these features for improved functional outcomes.

## Disclaimer

This device has not been approved or cleared as safe or effective by FDA. This device is limited by U.S. federal law to investigational use.

## Data Availability

All data produced in the present study are available upon reasonable request to the authors.

## Notes

### Competing Interest Statement

The authors have declared no competing interest.

### Funding Statement

No external funding was received.

### Author Declarations

IRB0779 and IRB0773 of Battelle Memorial Institute gave ethical approval for this work.

