## Supplementary for "Identifying alterations in hand movement coordination from chronic stroke survivors using a wearable high-density EMG sleeve"

### Inclusion and Exclusion Criteria

For physically impaired individuals, inclusion criteria address the minimum length of time since the stroke that led to the impairment. Inclusion criteria may also pertain to meeting dimensional requirements related to interacting with the system hardware (e.g., subject’s arm dimensions must be such that they can appropriately don an existing electrode sleeve design).

For populations with potential for cognitive impairment (e.g., stroke survivors), inclusion criteria indicating ability to follow 3-step commands and communicate verbally (e.g., at least able to provide yes/no responses with accuracy) apply.

Specific Inclusion Criteria include:

1. Males and females ≥ 18 years old

2. Chronic stroke survivors who are at least 180 days post-stroke

3. Ability to provide appropriate consent to partake in the study

4. Ability to follow 3-step commands and deemed by an occupational therapist to have the capacity to complete required upper extremity movements

5. Ability to secure transportation to attend scheduled study sessions

6. Stroke-related hand impairment that interferes with ability to complete activities of daily living and is classified as Stage 1-6 on the hand subscale of the Chedoke McMaster Stroke Assessment

Persons with life-supporting or sustaining equipment or critical non-removeable implanted electronic devices are excluded for safety reasons since it is not known if the experimental systems would interfere with this equipment.

Specific exclusion criteria include:

1. Presence of any other clinically significant medical comorbidity for which, in the judgment of the Investigator, participation in the study would pose a safety risk to the subject

2. Currently participating in physical rehabilitation (e.g., occupational or physical therapy) for stroke-related upper limb impairment

3. Co-occurring neurological condition (e.g., Parkinson’s disease, Multiple Sclerosis) or other neuromuscular disorder (e.g., Carpal Tunnel Syndrome, neuropathy) that, in the judgment of the Investigator, may influence study results

4. Individuals who are immunosuppressed, have conditions that typically result in becoming immunocompromised, taking chronic steroids, or currently receiving immunosuppressive therapy

5. Individuals having or requiring any of the following: implanted pacemaker, life supporting/sustaining equipment, or critical non-removable implantable electronic devices such as an insulin pump or neurostimulator. An implanted Medtronic LINQ monitor does not meet this criterion (i.e., patients with a LINQ monitor may participate in this study).

6. Persistent pain ≥ 7/10 in impaired upper extremity, as measured by Numeric Pain Rating Scale (0-10)

7. Individuals whose forearm is determined to be too small or too large to fit the electrode sleeve being investigated.

8. Individuals who are pregnant or plan to get pregnant during the course of the study (self- report).

### Figures

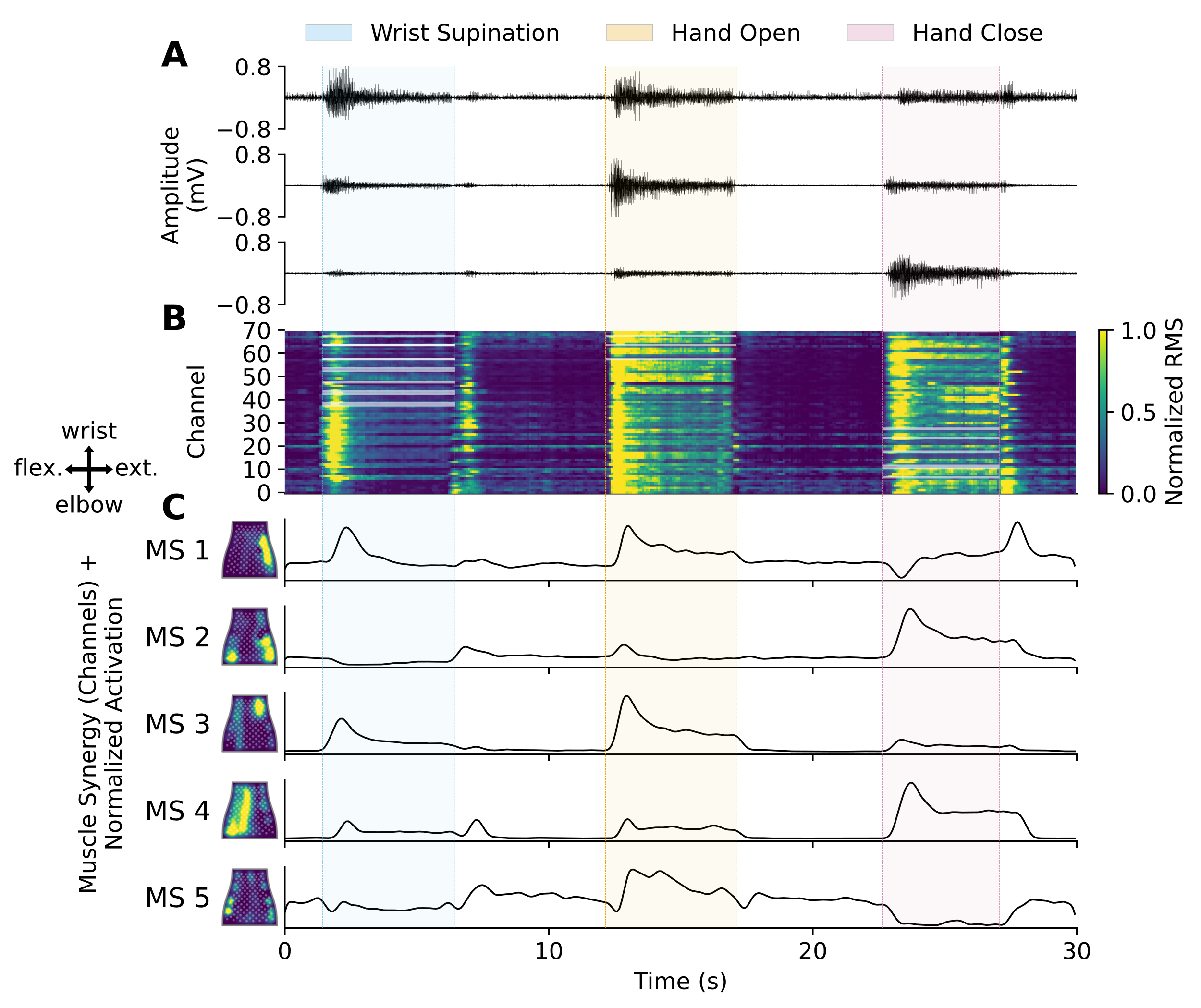
**Supplementary Figure 1. Representative EMG to HD-EMG feature decomposition pipeline across three cued movements from an able-bodied individual. (A)** Filtered EMG from three representative channels recorded using the NeuroLife sleeve. **(B)** Time-series heatmap of the globally normalized root mean square (RMS) of 150ms bins with stride of 40ms of EMG activity. Highlighted channels during the cued movements correspond to the agonist channels defined in the muscle group masks (Supplementary Figure 4 and Supplementary Table 3). **(C)** *Left:* Heatmaps of time-invariant muscle synergies decomposed with non-negative matrix factorization (NMF) mapped to the flattened EMG sleeve. Active areas correspond to the weighting of coupled muscles for each muscle synergy. A compass showing the orientation of the sleeve mapping is shown on the left (flex. = flexors, ext. = extensors). *Right:* Activation of the muscle synergies across time during the different attempted movements.

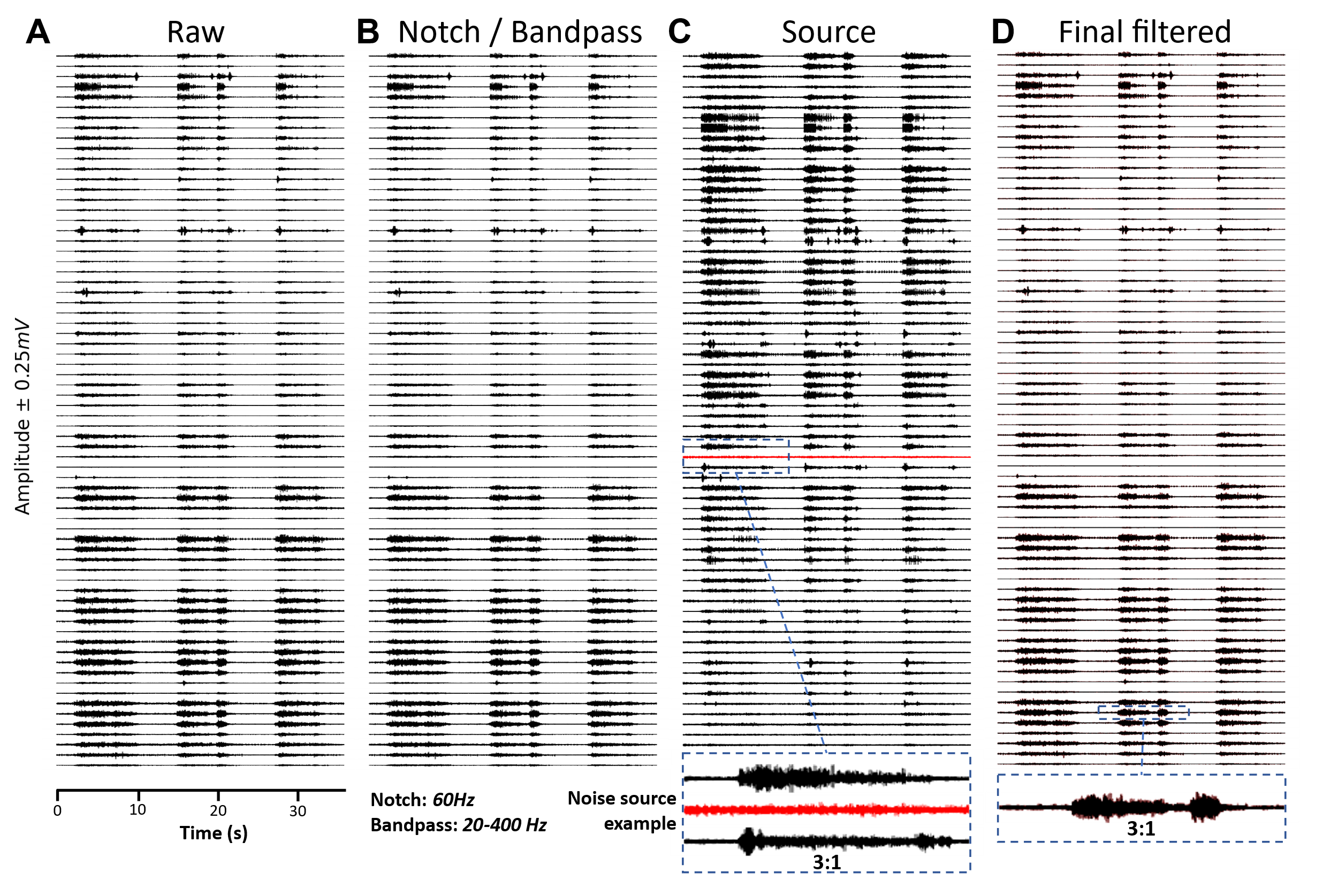

**Supplementary Figure 2. EMG filtering and artifact correction pipeline. (A)** Raw unfiltered EMG signal. **(B)** EMG signal after 60Hz notch filtering and 20-400Hz 10^th^ order Butterworth filtering. **(C)** Source activity from blind source separation through the approximate joint diagonalization of cospectral covariance matrices. The zoomed in region shows an outlier noise source to be removed from the reconstructed signal. **(D)** Reconstructed EMG signal with the outlier source suppressed. The zoomed in section shows a comparison between the signal in **(B)** (red) and **(D)** (black).

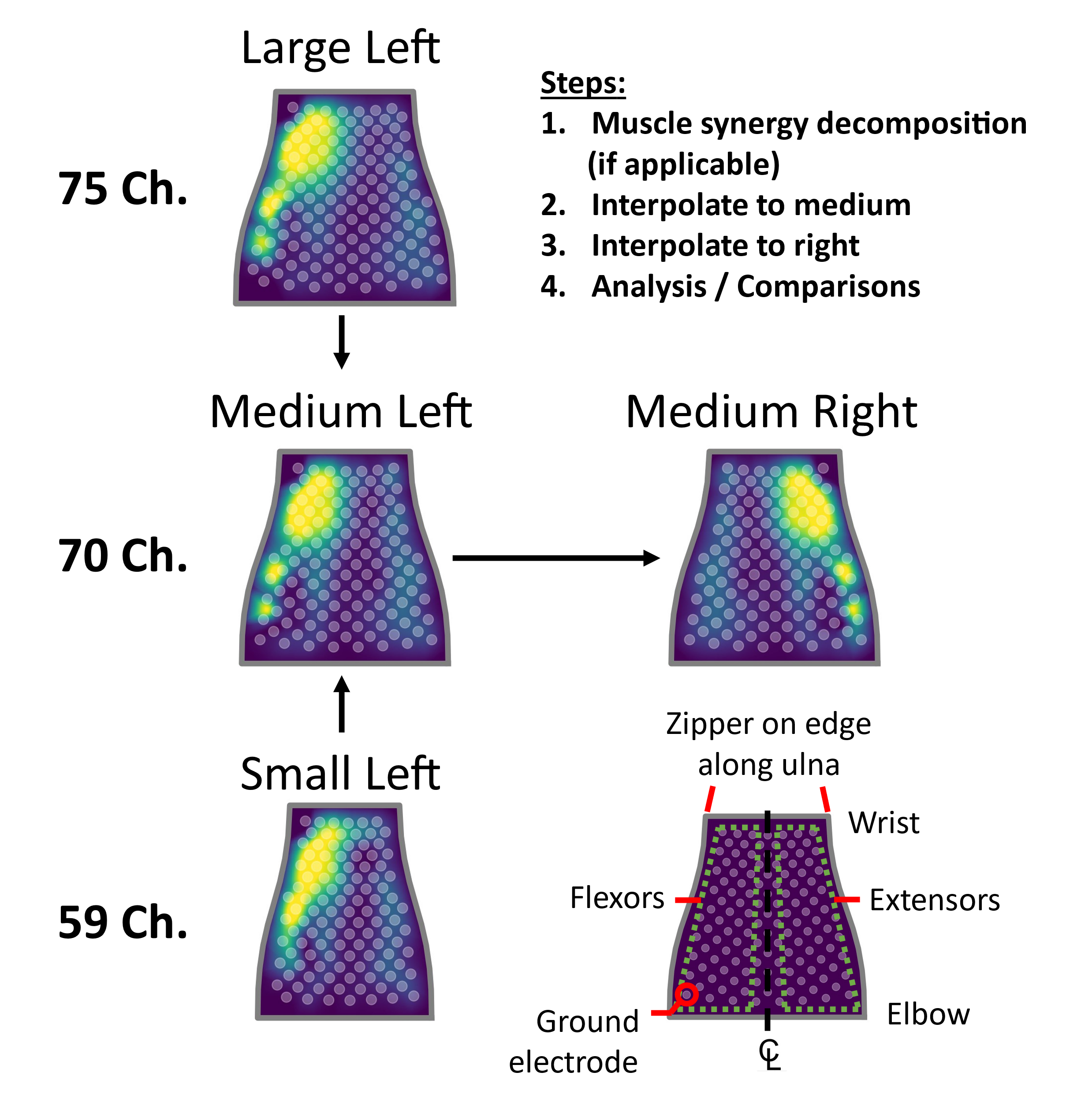

**Supplementary Figure 3. EMG sleeve mapping procedure.** All features and decomposed muscle synergies were mapped to the medium-right configuration. Once EMG RMS was computed from the sliding window EMG data, for co-contraction and muscle correlation analysis, the RMS data for all subjects was mapped to the medium sleeve. For muscle synergy analysis, muscle synergies were first decomposed into muscle synergies using non-negative matrix factorization (NMF) and then mapped to the medium sleeve. Following the sleeve size mapping, in the case the sleeve was worn on the left arm, the EMG feature date was mirrored to the right configuration. This ensured a consistent comparison between subjects.

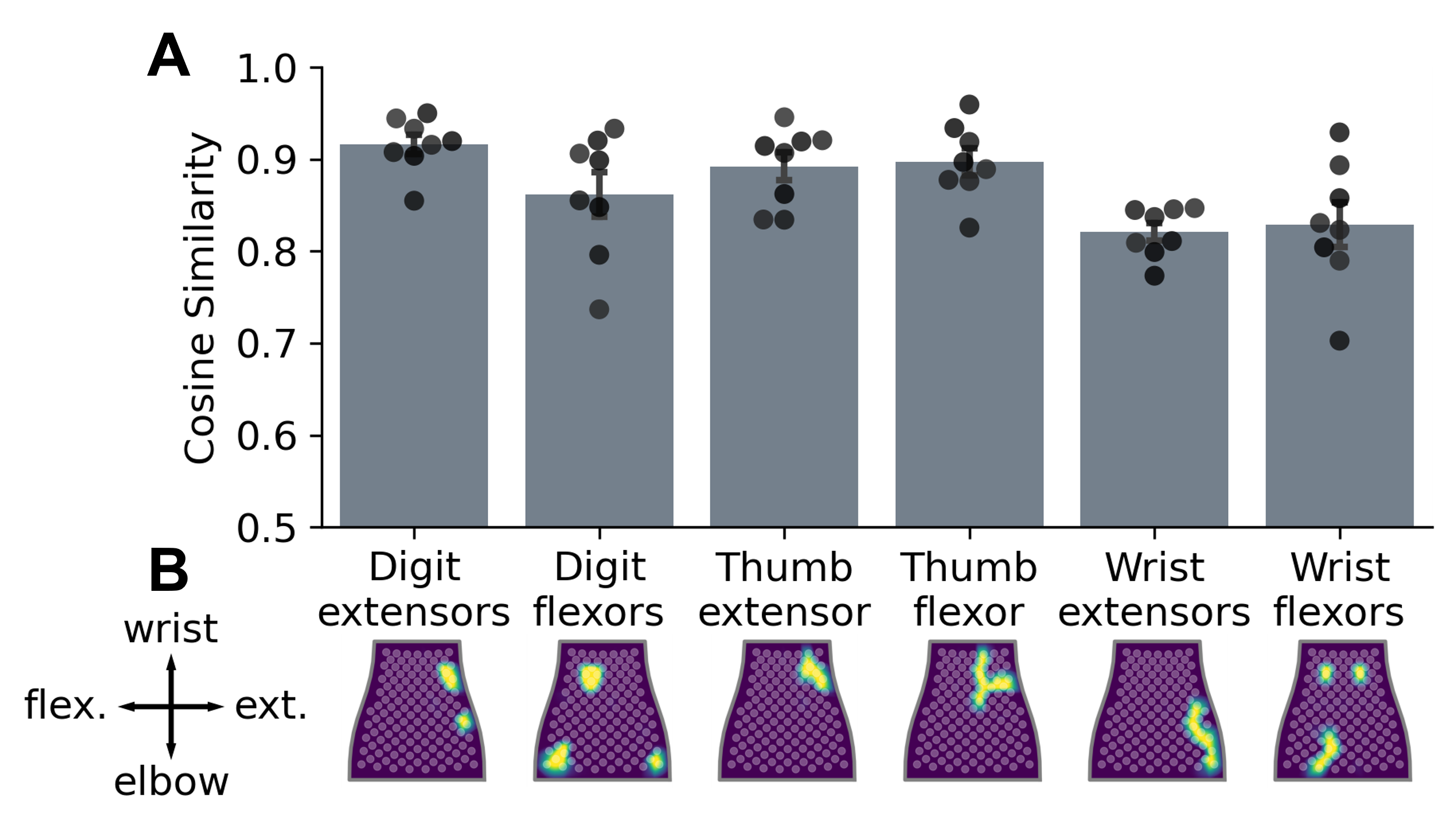

**Supplementary Figure 4. HD-EMG to muscle groups mapping analysis. (A)** Cosine similarity in a leave one out (LOO) comparison of muscle masks between each able-bodied subject and the remaining group average. **(B)** Muscle masks were generated from a 95% threshold of EMG activity during the associated movement and averaged within the session and across participants (Supplementary Table 3). Muscle masks were used to map the HD-EMG signal to muscle groups for physiological interpretation and to measure co-contraction index (CCI) and muscle correlations for different movements.

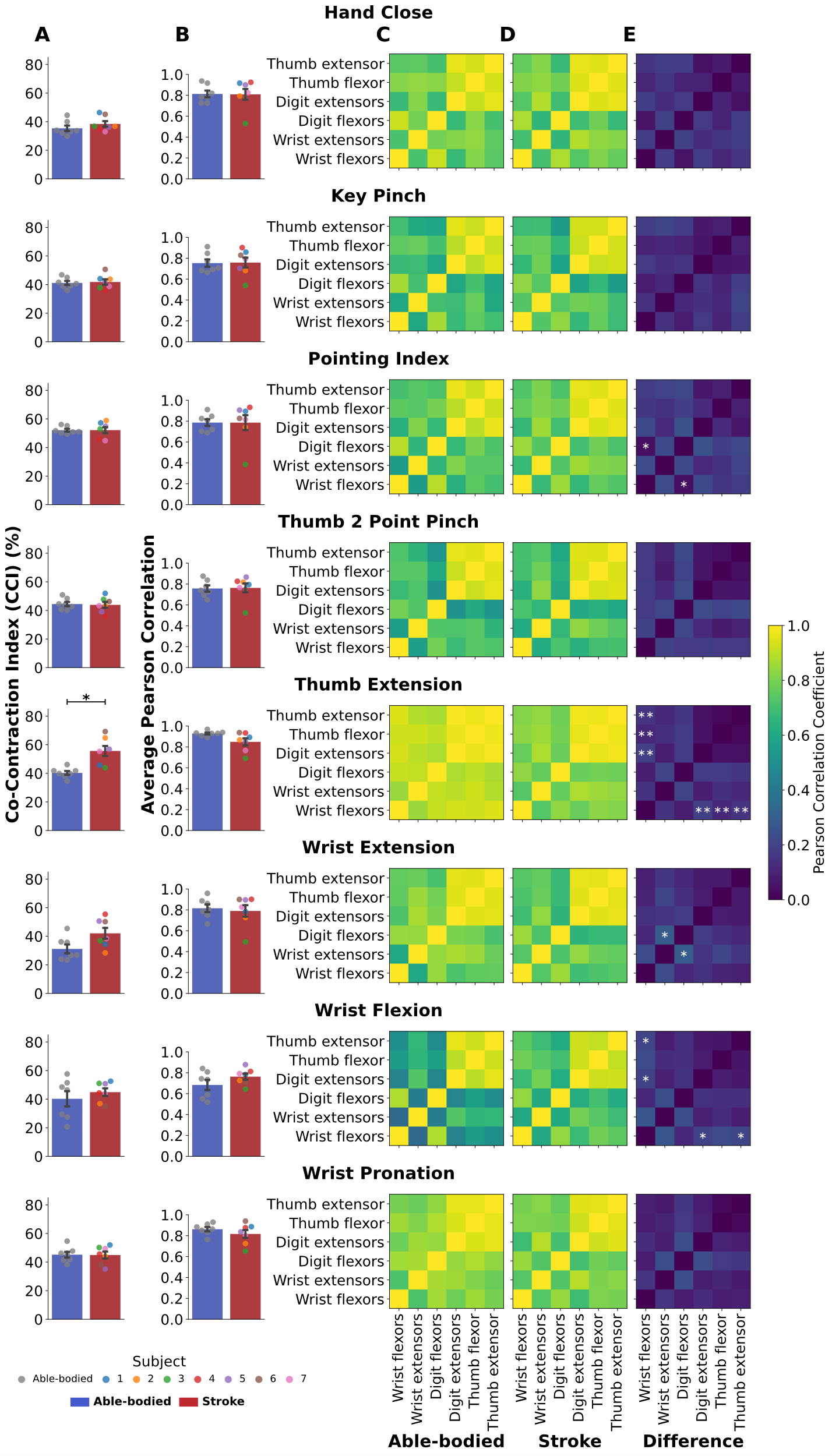

**Supplementary Figure 5. Co-contraction and muscle coupling across muscle groups for the remaining movements in the shared 12-movement dataset. (A)** Co-contraction index (CCI) and **(B)** average Pearson correlation between muscle groups for able-bodied (blue bars) and stroke (red bars) groups. Participants with stroke had a higher CCI for Thumb Extension than able-bodied subjects (p=0.014). **(C-E)** Correlation matrices between muscle groups across able-bodied, stroke, and the difference between the two groups show able-bodied subjects on average have more coupling between certain muscle groups when performing the movements. Coupled muscle groups determined with paired t-tests are denoted with * (p<0.05) and ** (p<0.01) indicating significant differences between groups.

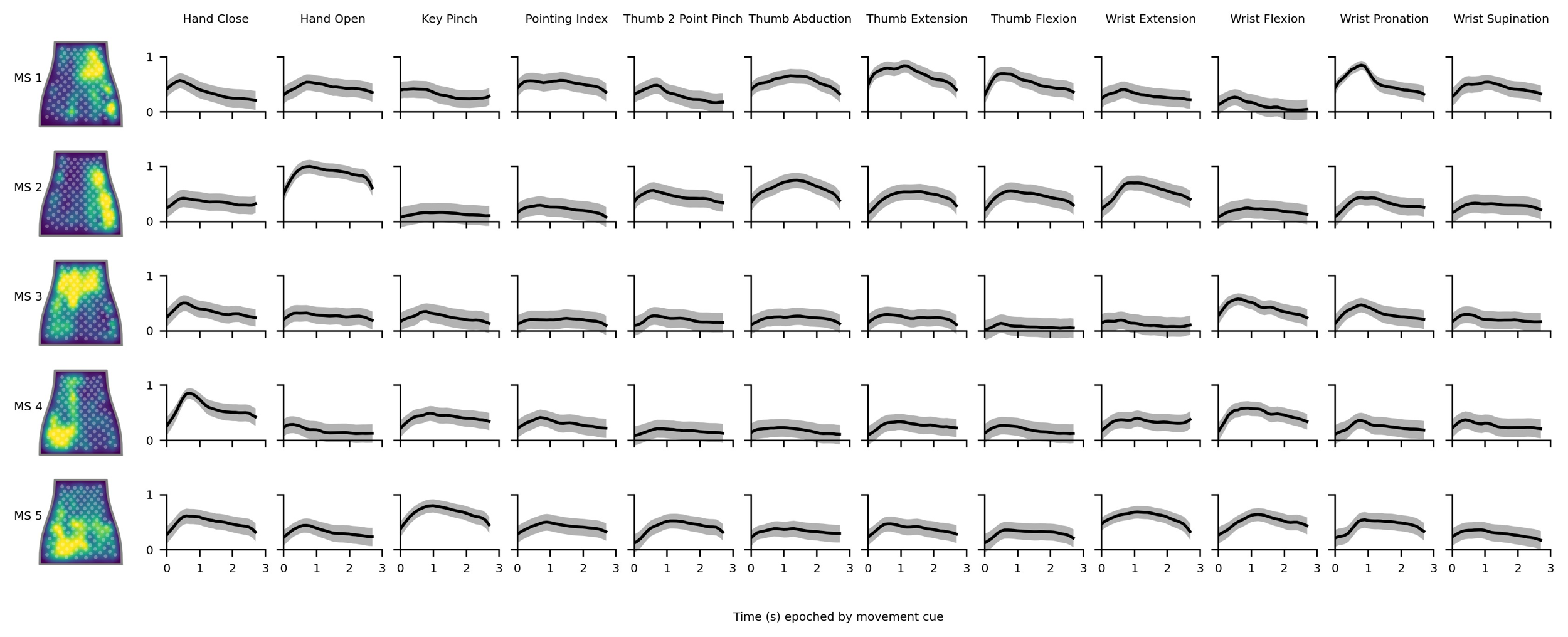

**Supplementary Figure 6. Average muscle synergies (MS) and synergy activations for participants with stroke.** *Left:* Time-invariant muscle synergies by channels mapped to the flattened NeuroLife sleeve. *Right:* Average synergy activations ± standard error of the mean across epoched movements including a 600ms cue shift to account for reaction time. There is a considerable amount of variation in the activation of synergies across movements.

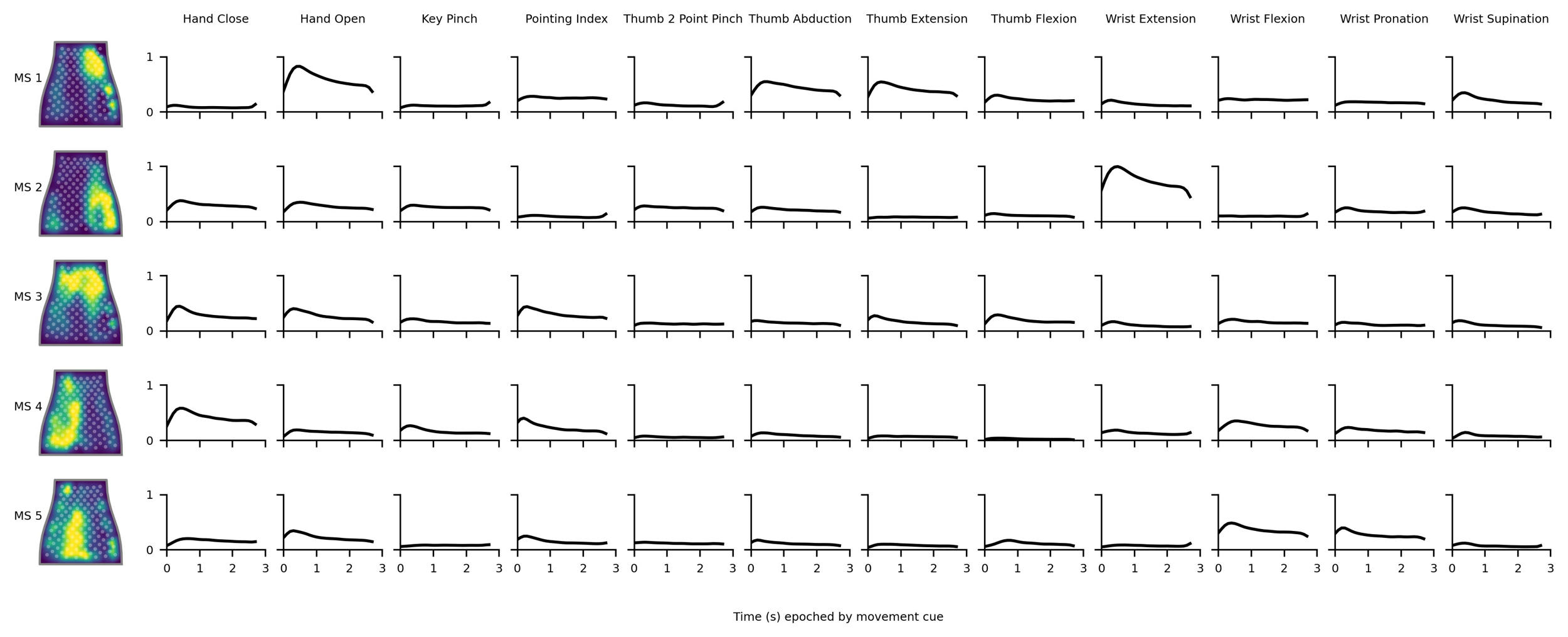

**Supplementary Figure 7. Average muscle synergies (MS) and synergy activations for able-bodied participants.** *Left:* Time-invariant muscle synergies by channels mapped to the flattened NeuroLife sleeve. *Right* Average synergy activations ± standard error of the mean across epoched movements including a 300ms cue shift to account for reaction time.

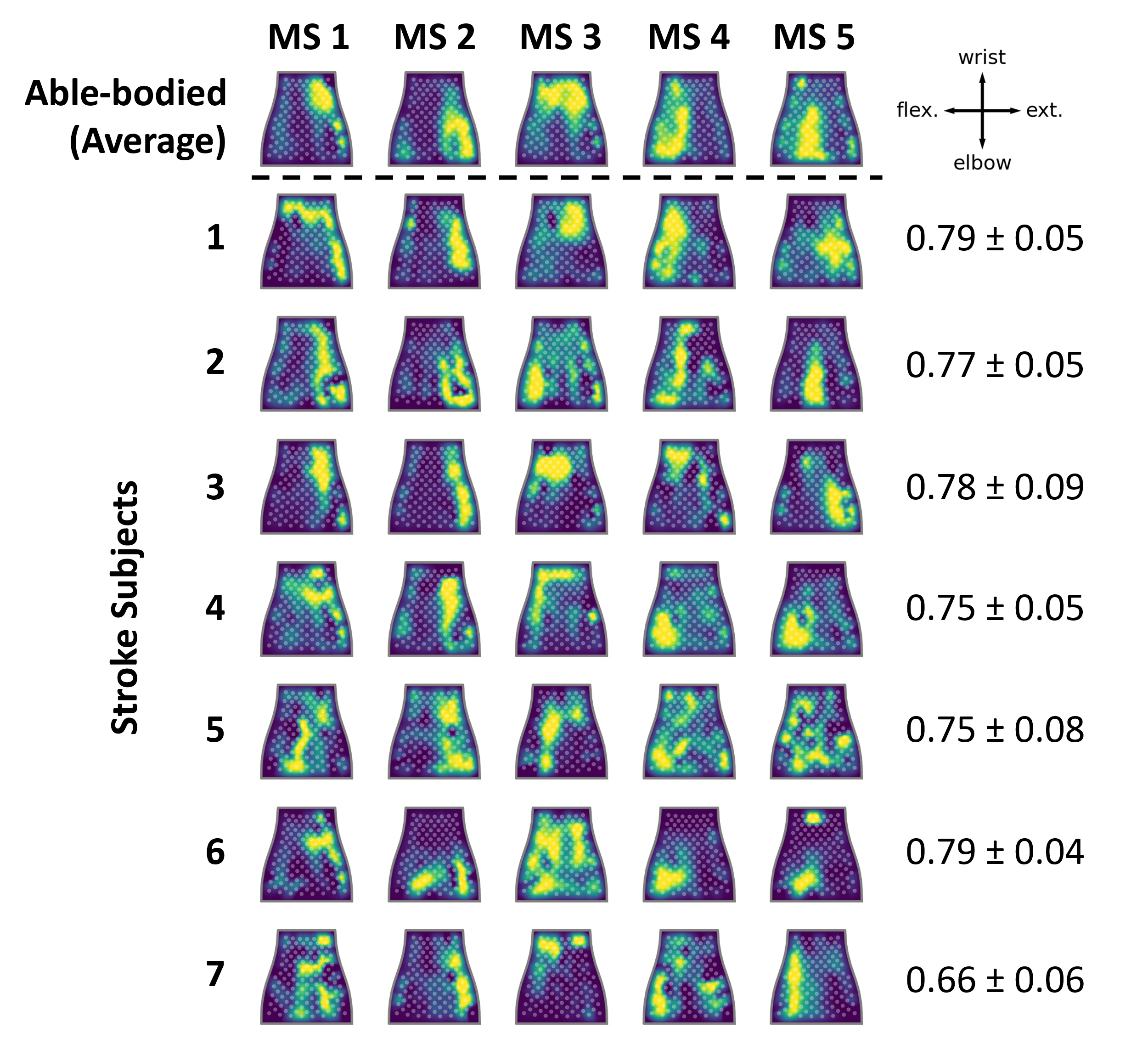

**Supplementary Figure 8. Comparison of muscle synergies (MS) by channels between participants with stroke and the average able-bodied group.** *Top:* Average time-invariant muscle synergies by channels mapped to the flattened NeuroLife sleeve. *Bottom:* Heatmaps of muscle synergies for each participant with stroke. *Right:* Cosine similarity ± standard error of the mean between stroke subjects’ muscle synergies and the able-bodied muscle synergy group average.

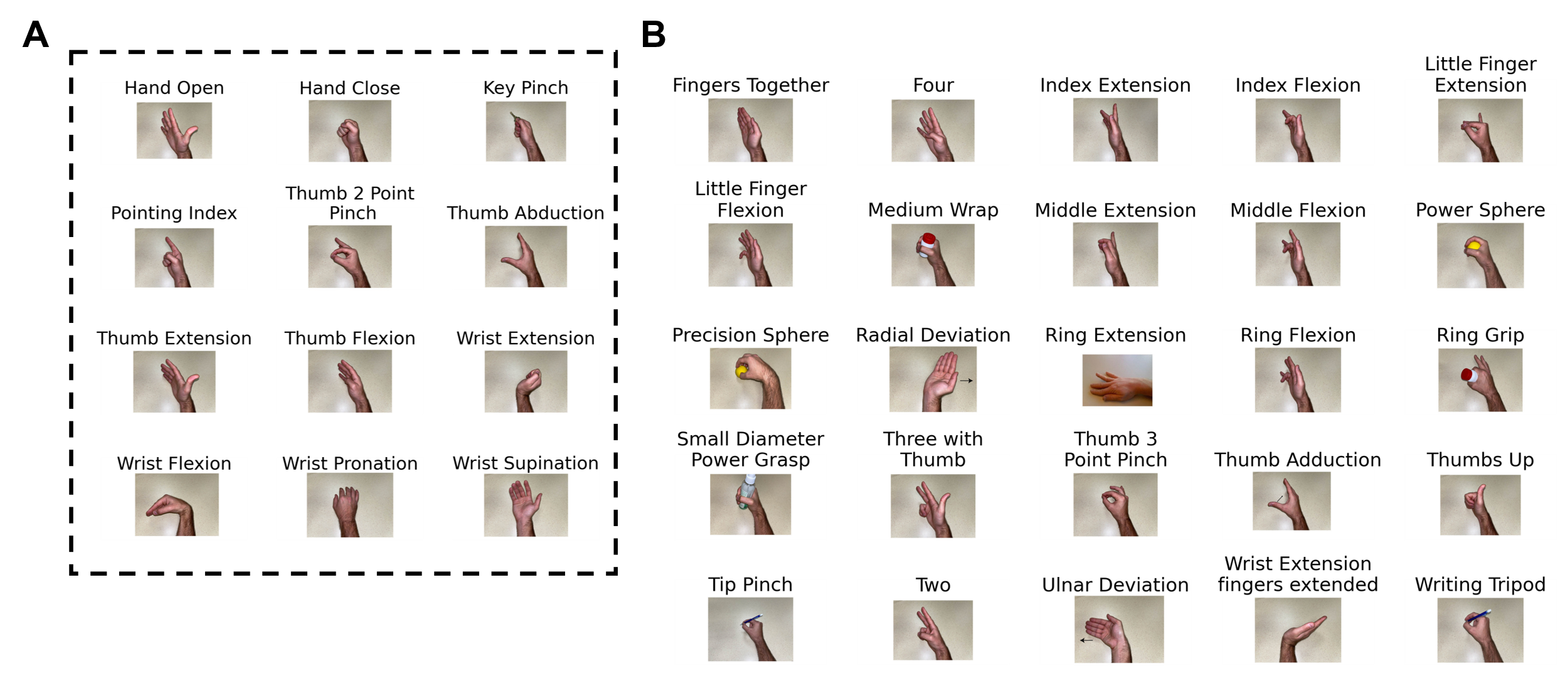

**Supplementary Figure 9. Pictorial cues for all movements from both datasets.** **(A)** 12-movement dataset that both stroke and able-bodied groups performed. **(B)** 25 additional movements for a total of 37 movements that able-bodied subjects performed for the benchmark dataset to assess movement coordination via muscle synergies.

### Tables

**Supplementary Table 1. Demographics, clinical metrics, and sleeve size of participants with stroke.**

| **Subject** | **UEFM** | **Time since stroke, years** | **Side of paresis** | **UEFM Hand** | **MAS Finger** | **MAS Wrist** | **Sleeve size** |
| --- | --- | --- | --- | --- | --- | --- | --- |
| 1 | 36 | 6 | Right | 6 | 1 | 1 | Medium |
| 2 | 22 | 4 | Right | 2 | 1 | 1 | Large |
| 3 | 32 | 3 | Left | 6 | 1 | 1 | Medium |
| 4 | 19 | 4 | Right | 4 | 1 | 0 | Large |
| 5 | 8 | 6 | Right | 0 | 4 | 3 | Medium |
| 6 | 7 | 7 | Right | 0 | 4 | 4 | Large |
| 7 | 38 | 6 | Right | 7 | 0 | 0 | Medium |

**Supplementary Table 2. Demographics and sleeve size of able-bodied participants.**

| **Participant** | **Age Range, years** | **Sex** | **Sleeve Size** |
| --- | --- | --- | --- |
| A1 | 26-30 | Male | Large |
| A2 | 31-35 | Female | Small |
| A3 | 26-30 | Female | Medium |
| A4 | 26-30 | Male | Large |
| A5 | 21-25 | Female | Small |
| A6 | 21-25 | Female | Medium |
| A7 | 26-30 | Male | Large |

**Supplementary Table 3. Movement and associated agonist muscle groups.** Italicized movements and muscle groups indicate movements from able-bodied participants that were used to generate muscle masks to map the HD-EMG sleeve to muscle groups (Supplementary Figure 4). All movements and agonist muscle groups were used for co-contraction index (CCI) calculation with the remaining muscle groups designated as non-agonist muscles.

| **Movement** | **Agonist Muscle Groups** |
| --- | --- |
| *Hand Open* | *Digit extensors* |
| *Hand Close* | *Digit flexors* |
| Key Pinch | Digit flexors, Thumb flexor |
| Pointing Index | Digit flexors, Digit extensors |
| Thumb 2 Point Pinch | Thumb flexor, Digit flexors |
| Thumb Abduction | Thumb extensor, Digit extensors |
| *Thumb Extension* | *Thumb extensor* |
| *Thumb Flexion* | *Thumb flexor* |
| *Wrist Extension* | *Wrist extensors* |
| *Wrist Flexion* | *Wrist flexors* |
| Wrist Pronation | Wrist flexors, Thumb flexor |
| Wrist Supination | Digit extensors, Thumb flexor, Thumb extensor |
